# Effect of a Pilates exercise program on the flexion-relaxation rate in women with chronic low back pain

**DOI:** 10.1101/2022.03.07.22270395

**Authors:** Ana Ferri-Caruana, Marco Romagnoli, Lluis Raimon Salazar-Bonet, Walter Staiano

## Abstract

**Purpose:** To evaluate the effect of a Pilates exercise program (PEP) on FRR and FRR asymmetry of the erector spinae (ES) muscle during standing maximal trunk flexion/extension in women with chronic low-back pain (LBP). A secondary goal was to investigate the effect of PEP on full trunk flexion ROM (TFRoM), pain intensity and functional capacity and analyse their relationship with the FRR.

**Material and methods:** Thirty women with chronic LPB were randomly assigned to either PEP (EG, n=15) or control group (CG=15). EG followed an 8-week PEP while no specific intervention was carried out on the controls. Before and after this period all variables were recorded.

**Results:** FRR did not show any significant changes between or within groups (p>0.05). EG showed a significant statistical difference in the FRR asymmetry pre- and post-intervention (p□ 0.05). Full TFRoM did not show any significant changes between or within groups (p>0.05). EG showed a significant decrease of 30% on pain intensity and a significant increase of 13.4 % in functional capacity (P□0.001) from pre to post-intervention.

**Conclusions:** An 8-week PEP does not affect FRR nor full trunk ROM, however yields improvements in pain intensity and functional capacity.Professionals should be aware of the negative effect on FRR asymmetry.

## Introduction

Clinical practice guidelines recommend exercise therapy for patients with chronic LBP [1,2] with the goal of improving disability and reducing absence from work due to physical and functional recovery [3,4]. In this regard, Pilates is not only widely extended in social physical activity programs and recommended among health care professionals [5] but also it has been increasingly incorporated into physiotherapy rehabilitation programmes [6-10] for those with chronic LBP.

Pilates exercises are performed based on five traditional principles: centering, control, concentration, breath, precision, and flow. These principles unite the body and the mind, and may affect positively performance of the different exercises (i.e. deep breathing, attentional internal focus) and therefore have a positive influence on EMG parameters of lumbo-pelvic muscles [11-13]. For instance, Silva et al. [13] showed that Pilates exercises *double leg stretch, coordination, crisscross* and *foot work* promoted greater muscle activation than traditional exercises in the upper rectus abdominis.

One consistent finding in patients with LBP is an ability to display the flexion-relaxation phenomenon (FRP) during full trunk flexion. The FRP is defined as reduced activity of lumbar extensor muscles in standing maximum trunk flexion [14].

The flexo-relaxation ratio (FRR) is a measure for quantifying FRP and is defined as the ratio of the peak EMG amplitude during the flexion motion to the EMG amplitude in full flexion. A smaller flexion-relaxation ratio (FRR) [15,16] and a smaller extension-flexion ratio (EFR) [17] in patients with back pain are the most common differences in ES muscle activity as compared with healthy subjects, indicating that neuromuscular coordination between the trunk and the hip could be abnormal in patients with CLBP. The literature review carried out by Colloca et al. [18] concluded that not only the FRP assessment procedure was reliable but also showed that the absence of the FRP in patients with chronic LBP could be corrected with treatment.

There is some controversy regarding the effect of some exercise therapies on the FRP. For instance, conventional physical therapy or a lumbar stabilization exercise program have shown no effect on FRP [19-21] . On the contrary, other studies have shown an increased FRR after a 12-week lumbar spinal exercise program [16,22] or a higher number of patients with a normal flexion-relaxation level and lumbar ROM after a functional restoration rehabilitation program [23].

Furthermore, previous studies have investigated asymmetry in the FRP [24-26] since as previously mentioned an imbalance in trunk muscle activation between the right and left sides can induce pain intensity by loading the spine incorrectly in patients with non-specific chronic LBP [27,28].

Shahvarpour et al. [21] tested the effect of an 8-week lumbar stabilization program in people with chronic LBP on the EMG/kinematics of paraspinal muscles (longissimus, iliocostalis and multifidus) during trunk maximal flexion-extension. Although they found positive changes on pain intensity and disability following the intervention, the FRR was not changed concomitantly. However, there was not a group control; thus there was no way to compare results with paired participants. FRR asymmetry measures were also not considered.

Therefore, the aim of this study was to evaluate the effect of an 8-week Pilates exercise program (PEP) on FRR and FRR asymmetry of the erector spinae (ES) muscle during standing maximal trunk flexion-extension in women with chronic LBP. It was hypothesized that the PEP would increase the FRR and would decrease the FRR asymmetry. A secondary aim of the study was to determine the effect of the PEP on clinical outcomes such as full TFRoM, pain intensity and functional capacity in women with LBP, and explore the relationship with the FRR.

## 2. Material and methods

### 2.1 Participants

The sample size was determined by preliminary power analysis to avoid 5% -error and 95% power of the text. Sample size of 15 subjects in each group was obtained. The sample size was increased to account for possible dropouts.

Participants were aged between 45 and 65 years old, and were recruited from an advertisement in a community sports halls where many physical activities were delivered (i.e. Pilates, yoga, aerobics, TRX, …). Seventy-nine people who joined the Pilates activity were contacted before the beginning of the classes. Fifty-nine people were interested in participating in the study and completed the Oswestry Disability Index. Finally, sixteen participants met the inclusion criteria and were included in the experimental group (EG). Posteriously, from a convenient sample (referrals from the EG), sixteen women with similar characteristics and lifestyle, met the inclusion criteria and were recruited as a control group (CG). There were two drop-outs, one from the EG (abandoned PEP because of a lower leg related injury) and one from the CG (received back physiotherapeutic treatment). Therefore, the final number of participants for each group was 15.

The inclusion criteria were: lumbar or lumbosacral pain (with or without radicular pain) for at least six months; a score higher than 6/50 on the Oswestry Disability Index (ODI) [29.30] and absence of any back treatment for the last three months.

The exclusion criteria were: body mass index > 30 kg/m2; prior surgery of the pelvis, spinal column or lower extremity; scoliosis; systemic or degenerative disease; history of neurological diseases or deficits not related to back pain; pregnancy or hypertension. Before testing, each participant was informed of all experimental procedures and provided their informed written consent. The ethics committee of the University of V. approved the study and the consent form. The study was conducted in compliance with the Declaration of Helsinki.

### 2.2. Exercise intervention

The PEP (1 h x 2 sessions per week x 2 months) was delivered by one Pilate’s professional (with a mean of 6.5 years of experience in Pilates). The PEP carried out by Pilate’s instructor has been published in a previous study [31]. The exercises focused on core stability, posture, breathing, flexibility, strength, and muscle control, being the active awareness of the use of trunk muscles to stabilize the pelvic-lumbar region the main approach. The exercises performed were: the hundred, the rolled up, single leg circles with bent leg, spine stretch, rolling like a ball and single leg stretch. Each exercise was performed as follows: 4 repetitions of 30 seconds with 2 minutes of recovery between repetitions. In order to complete the 60 minutes session two exercises: the “superman” and the double leg bridge were added to the mentioned PEP.

Before the start of the exercise program, all participants from EG received a 1-hour basic introduction to the PEP and were trained in how to activate the core muscles, which involve isometric contraction of the transversus abdominis, pelvic floor, and multifidus muscles while exhaling during diaphragmatic breathing. Adverse events were monitored by pain intensity during the execution of the exercises and before and after sessions. All exercises were performed on a rubber mat of minimum ¾ inch thick.

The attendance was weekly recorded for each participant. No cointervention was allowed for any of the participants in the study. Both groups continued their usual lifestyle and were allowed to use NSAIDS only if needed. Every three days telephone calls were made to all participants in order to assure that any unusual physical activities were performed.

### 2.3. Questionnaires

The measure used for LBP-related disability was the transcultural adaptation to the Spanish population of the ODI [29,32] . This questionnaire was used as inclusion criteria. Subjects could participate in the study if they obtained a minimal score of 12% [28].

The version adapted to the Spanish population of the Low Back Outcome Score (LBOS) questionnaire [33,34] was used to measure self-reported functional capacity. A 10-cm Visual Analog Scale (VAS) was used to evaluate pain intensity during the last week [35,36]. The left end of the line was anchored with “no pain,” and the right end of the line was anchored with “worst pain”.

LBOS and VAS were measured at pre-intervention (T0) and pos-intervention (T8).

### 2.4. Full trunk flexion ROM

To obtain the full trunk flexion ROM (TFRoM), the angular position of the inertial EMG sensor (attached with a kinesio-tape at T3 vertebrae) was measured from the start to the end of the flexion movement. Full TFRoM was calculated as the average TFRoM from the three flexion-extension tasks.

### 2.5. Flexion-extension tasks

All subjects were assessed with the flexion-extension task pre and post control and intervention periods (T0 and T8, respectively).

The subjects stood with their arms by their side and their feet shoulder width apart. Beginning in standing, the subjects were asked to: bend forward as far as possible (4 s to flex); relax in the fully-flexed position (4 s to relax); return to upright standing (4 s to extend); stand quietly for 4 s. This was repeated 3 times in succession. A metronome was used to pace the movements. Subjects were instructed to keep their knees straight, to not contract the abdominal muscles, and to keep their head fully flexed to minimize cervical movement.

### 2.6. EMG measurements

The electromyographic activity was recorded from the ES right and left muscles with one portable 2-channels device from the Shimmer branch (Realtime Technologies Ltd, Dublin, Ireland) with a 16-bit analog / digital (A / D) conversion. The sampling frequency was programmed at 1024 Hz.

During the registration, the EMG signal was monitored using the mDurance software (MDurance Solutions S.L., Granada, Spain) for Android and stored in a cloud server for further analysis.

The mDurance software digitally filtered the raw signals automatically through a “Butterworth” band pass filter of the fourth order between 20 and 450 Hz. A cutting frequency for the high-pass of 20 Hz was used to reduce the “artifacts” that could arise during the movement to have an impact minimum in the total power recorded by the EMG [37].

The root mean square (RMS) was calculated from the filtered EMG signal. The mean RMS values, measured during the time that the different tests lasted in each of their movement phases were used for the analysis.

The skin was shaved, rubbed, and cleaned with alcohol. Bipolar pre gelled electrodes Ag/AgCl surface electrodes (MedCaT B.V, Doorndistel, Spain) were used to record the EMG. Electrode placement were as SENIAM guidelines indicate [38].

Asymmetry in muscle activity between both (right and left) ES muscles was calculated as their mean difference in RMS in the three flexion-extension tasks.

FRR was calculated by dividing the maximal EMG amplitude during flexion by the minimum EMG amplitude at full flexion [15,22]. The mean of the 3 trials performed was used to determine the FRR for each muscle for each subject. A lower FRR indicated a greater state of muscle relaxation.

Asymmetry in the FRR was calculated as the absolute difference between FRR of the right and left ES muscles.

### 2.7. Statistical analyses

The SPSS software (version 21; SPSS Inc., Chicago, IL) was used for the statistical analysis.All data are presented as mean and standard deviation (SD), unless otherwise stated. Assumptions of statistical tests such as normal distribution and homogeneity of data were checked with the Shapiro-Wilk test and Levene test, respectively.

Independent T-tests were carried out to measure differences at baseline. Two-way repeated measures ANOVA (GROUP X TIME) were carried out for all dependent variables. Significant interactions or main effects were further analysed using a post-hoc Tukey-Kramer test. The effect sizes for repeated measure ANOVA were calculated as partial eta squared (n^2^p) using the small= 0.02, medium= 0.13 and large= 0.26 interpretation for effect size. For pain, functional capacity, full back ROM and ES EMG variables correlational analysis (Pearson’s correlations) were carried out for all subjects at baseline. For each ES EMG variable correlational analyses (Pearson’s correlations) were carried out in participants with LBP to test for the “potential” (hypothesis generation stage) presence of moderator effects or of mechanisms operating during the treatment. A p value of .05 was accepted as the level of significance for all statistical analyses.

## Results

### Participants

The age, height, weight and BMI were not significantly different between groups at baseline. There were no differences at baseline in the variables of the FRR (p =.708), FRR asymmetry (p =.973), full TFRoM (p =.450), pain intensity and functional capacity. There were no significant differences between groups at baseline in the LBP disability index ODI (p =.223), the mean score for CG and EG was of 15.4 % and 10.2 % respectively, higher than the minimal score allowed to participate in the study (14%, equivalent to 7/50). See Table 1.

**Table 1.** Mean and SD of subjects’ anthropometric measurements, functional capacity and pain status.

| | CG (n=15)<br>X ± SD | EG (n=15)<br>X ± SD | *p-values* |
| --- | --- | --- | --- |
| Age (years) | 56.1 ± 5.4 | 55.5 ± 4.6 | 0.74 |
| Height (cm) | 157.4 ± 1.5 | 161.5 ± 0.9 | 0.86 |
| Weight (kg) | 66.9 ± 6.0 | 63.7 ± 5.6 | 0.14 |
| BMI (kg/m<sup>2</sup>) | 26.0 ± 2.0 | 24.7 ± 3.0 | 0.18 |
| ODI (/50)) | 13.7 ± 4.4 | 11.1 ± 4.5 | - |
| VAS (/10) | 5.9 ± 1.8 | 5.8 ± 1.8 | 0.93 |
| LBOS (/75) | 46.9 ± 7.4 | 47.9 ± 6.8 | 0.67 |
| LBP prior to study (weeks) | 18.8 ± 6.3 | 19.7 ± 7.5 | 0.34 |

\*n=number of subjects; X=mean; SD= standard deviation; BMI= body mass index; cm= centimetre; kg= kilogram; ODI= Oswestry disability index; VAS= Visual analogue scale; LBOS=Low Back Outcome Score; LBP=Low back pain.

None of the 15 participants missed more than 1 session as required to be included in the analysis.

### Effect of the Pilates exercise program on FRR and FRR asymmetry

There was a significant GROUP x TIME interaction for the variable of FRR asymmetry (p= .044, η^2^= 0.137) (Fig 1). Follow-up tests revealed that the EG showed a significant (p=.033) increase in FRR asymmetry at post test compared to baseline, while no significant differences were found in the CG from baseline to post test.

**Figure 1.**
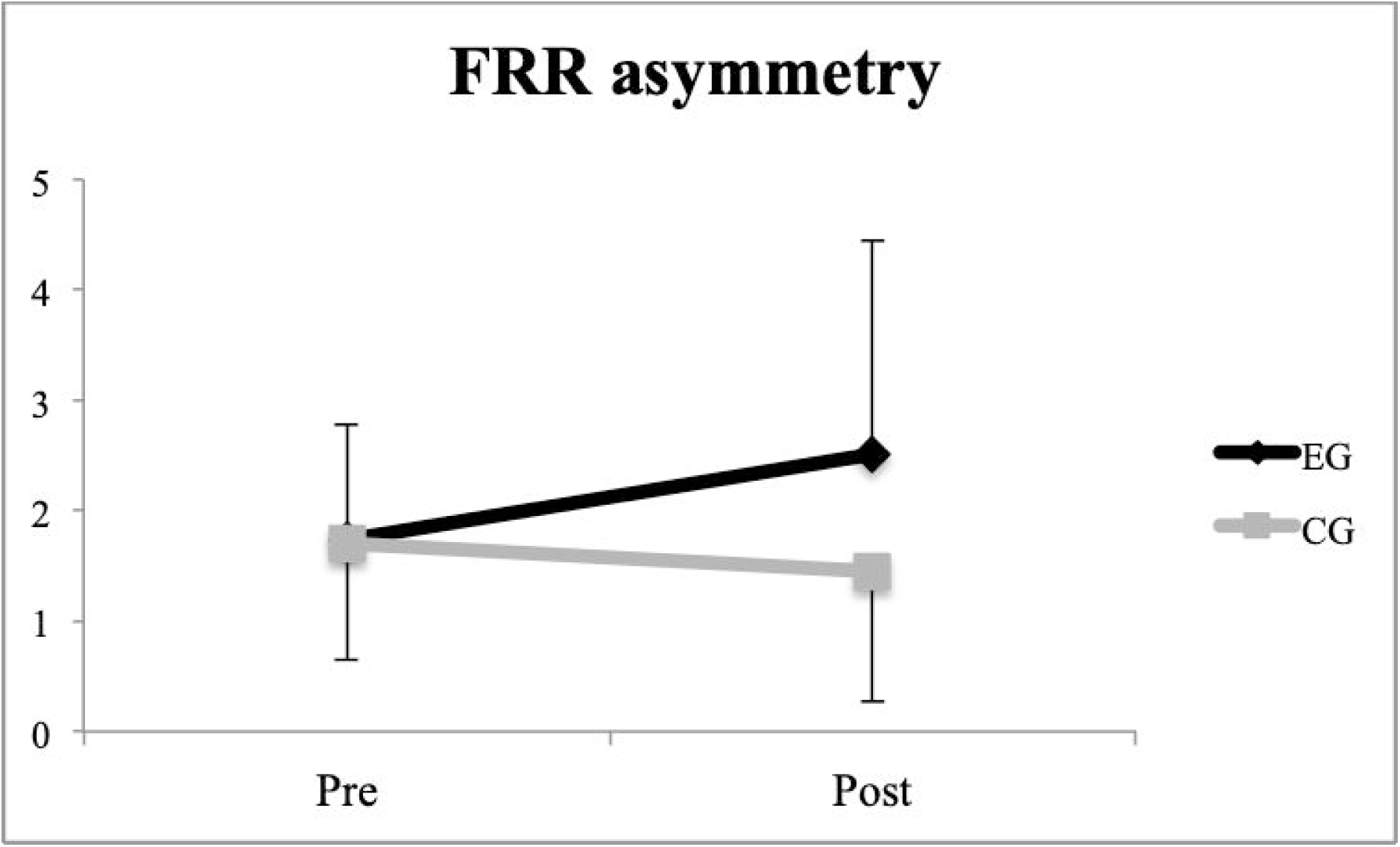
FRR asymmetry for each group at pre and post-intervention.

### Effect of the Pilates exercise program on full trunk flexion range of motion, pain intensity and functional capacity and their relationship with FRR

Full TFRoM did not show any significant changes between or within groups (p>0.05). There was a significant GROUP x TIME interaction for the variable of pain intensity (p= 0.003, η^2^= 0.268) (Fig 2). Follow up tests revealed that participants in the EG rated the pain intensity significantly lower (30%) at post-test compared to baseline, while participants in the CG did not show any significant difference in the pain intensity rating from baseline to post interventions.

**Figure 2.**
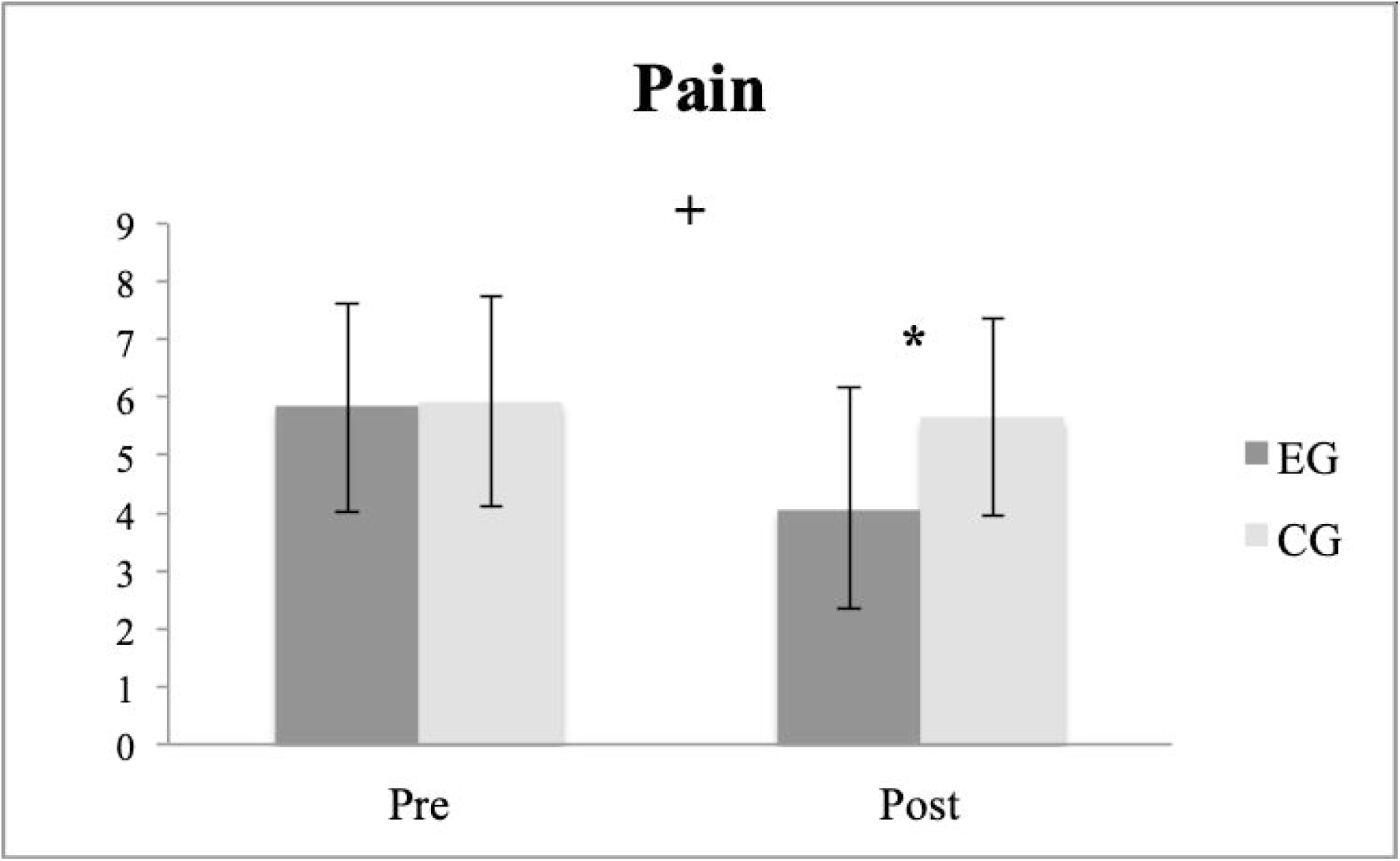
Numeric pain rating scale for each group at pre and post-intervention. (mean ± SD), * p □ 0.05, + p □ 0.05.

There was a significant GROUP x TIME interaction for functional capacity (p□ .001, η^2^= 0.570) (Fig 3). Follow-up tests revealed that the EG significantly (p□ .001) increased functional capacity (13.4%) at post-test after the PEP intervention, while no significant difference was reported for functional capacity in the CG.

**Figure 3.**
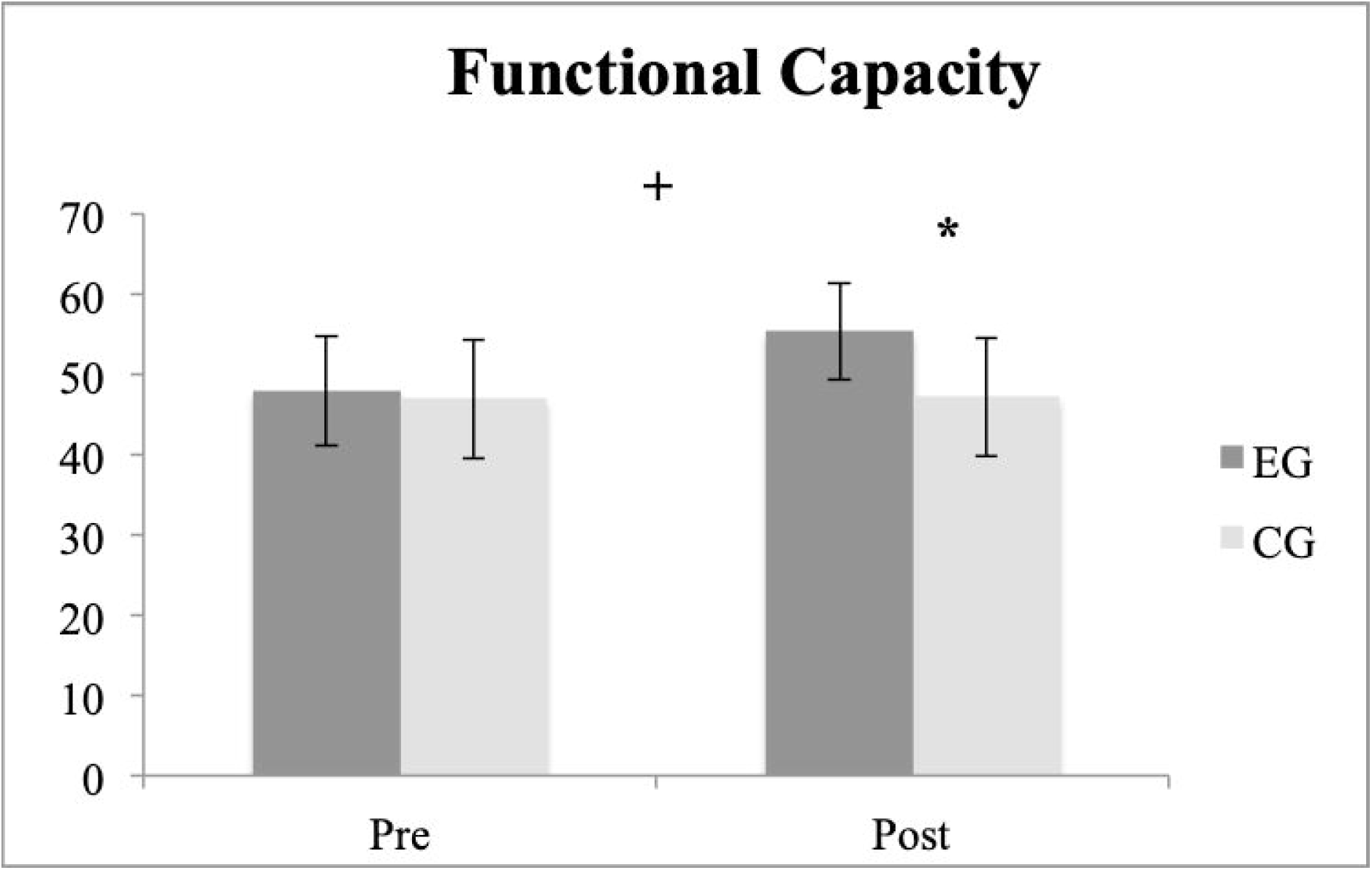
Functional capacity rating scale for each group at pre- and post-intervention. (mean ± SD), * p □ 0.001, + p □ 0.001.

FRR did not show any significant interactions or main effects with full TFRoM, pain intensity and functional capacity. But, correlational analysis showed that FRR was positively but weakly correlated with functional capacity (r=0.40, p□ 0.05).

## Discussion

The aim of the present study was to evaluate the effect of an 8-week PEP on FRR and FRR asymmetry of the ES muscle during standing maximal trunk flexion/extension in women with chronic LBP. A secondary goal was to investigate the effect of the PEP on full TFRoM, pain intensity and functional capacity and their relationship with FRR.

Since Pilates has shown positive results on EMG parameters due to its principles when compared to other studies using conventional therapy or strength and stability exercises in chronic LBP patients [19-21], we expected to find an improvement in the FRR. However, our results did not show any significant changes between or within groups.

As in our study, Shahvarpour et al. [21] did not observe a FRR after an 8-week stabilization program in a group of chronic LBP patients. This could be explained by the fact that in our study participants had no previous Pilates experience. Performing basic Pilates exercises imply learning new concepts such as breathing, concentration and attentional focus while performing an exercise and this *mental* part might be a challenge for beginners. A longer familiarisation/intervention period or doing the study on experienced participants may have provided more positive results on FRR.

Our results differ from other studies. Neblett et al. found that 94% of those chronic LBP participants concluding the treatment achieved FRP, even if several failed to improve ROM to normal levels. However, they used a sEMG biofeedback training to teach patients how to relax their backs during flexion, which may have positively influenced the results of the dynamic test of full trunk flexion-extensión movement. Marshall et al. [22] also found an improvement of the FRR in chronic LBP patients, however, there was no way of verifying if the patients showed an impaired FRP at baseline or any changes in PRP following the intervention, since there was no healthy control group.

FRR asymmetry was statistically significantly higher from pre- to post intervention. (p <0.05). As studies on FRR asymmetry after an exercise intervention program are scarce, comparison with other studies is difficult. However, since the focus of the PEP program is deep muscle strengthening in a symmetrical fashion and correct alignment of the trunk, we expected an improvement on FRR asymmetry. In this line, Rutkowska et al. [40] did not measure the FRR asymmetry, but they found that asymmetry of EMG activity in ES after a 4-week training programme was maintained yet it was not statistically significant. This is the first study showing that a PEP program has a negative effect on FRR asymmetry. A possible explanation may be due to the fact that the PEP participants start the program with LBP and they may be unconsciously altering the movement pattern of exercises to avoid pain. So, health care professionals should be aware of possible negative motor control adjustments of LBP participants during a PEP program. We did not specify participants LBP location (right-left), therefore it would be interesting for future studies to determine the relationship between LBP location and FRR asymmetry after an exercise intervention.

More research on EMG parameters is necessary to give scientific support to the philosophical and conceptual premises of Pilates if it is to be systematically included in rehabilitation and training programmes.

There is controversy regarding the effect of a Pilates exercise intervention on trunk ROM in people with chronic LBP [21,41-43] . The results of Shahvarpour et al. [21] are very similar to ours since they found that although pain was decreased and functional capacity improved, full TFRoM was not improved concomitantly. Our study supports the idea that not only TFRoM does not change but FRR does as well. This could be explained by the fact that participants from the study presented a low score in the ODI (11/50), thus changes in neuromuscular efficiency improvement may be more difficult to achieve. Further research should investigate whether similar findings exist in individuals with more severe pain or higher levels of disability.

Our results on pain reduction are in line with most of the studies that show improvements (between 16% - 54%) on pain after a PEP intervention [6-8,41-46]. Therefore, we support the idea that Pilates exercises are a good option to improve pain intensity, but our study shows no relationship between pain improvement and FRR. Our hypothesis is that participants with chronic LBP may have learnt to stiffen the lumbar spine during the PEP, producing no changes on FRR during the trunk flexion-extension task.

Regarding functional capacity, our participants improved 9.8 %, which is a result lower compared to the result obtained by Kofotolis et al. [47] with a 33.3% improvement on disability in female chronic LBP patients. Improvement on function in the present study (7.4 points) can be considered clinically significant, as a improvement of 7.5 (10%) change in the 75 point LBOS is widely used as a criterion of clinically significant change for patients with only LBP [48]. Interestingly, FRR was positively correlated with functional capacity. One previous study [49] has demonstrated that increases in the FRP following a 12-week exercise intervention was the best predictor of improvements in self-reported disability. However, in our study the correlation of functional capacity with FRR was very weak, therefore we cannot assume that the improvement in functional capacity is explained by an improvement in the FRR.

This study had some limitations. First, the fact that patients were recruited from advertisements, may affect the generalizability of results. Secondly, using one inclinometer to measure TFRoM does not account for movements of the pelvis. Methods using twin inclinometers or the motion system analysis could be alternatives, as these would remove any pelvis movement from the equation.

## Conclusion

The intervention of an 8-week PEP in women with chronic LBP did not positively or negatively affect the FRR, but it did negatively affect the FRR asymmetry (primary outcome). Health professionals should be aware of the increased asymmetry on the ES that is caused by the application of a PEP program on chronic LBP patients. Further research is needed to understand the cause of pain intensity and functional capacity improvement after a PEP program.

Furthermore, the PEP yielded improvements in pain intensity and functional capacity (secondary outcome) and it can be administered safely since it is well tolerated by female chronic LBP patients.

## Data Availability

All data produced in the present work is going to be submited with this manuscript.
Raw data are available upon reasonable request to the authors.

## Acknowledgements

The authors would like to thank Berta Dabaliña, Amparo Soler and the professional staff from “Sagunto Patronato Municipal de Deportes” for participant recruitment and delivery of the PEP program.

Financial Disclosure statement: The author(s) received no specific funding for this work.

